# Prolonged Pain in Premature Neonates Hospitalised in the NICU: A Principle Based Concept Analysis

**DOI:** 10.1101/2025.09.03.25335043

**Authors:** Alexandra Breton-Piette, Bianca Breton-Piette, Abir Rebhi, Clémence Coupat, Marilyn Aita

## Abstract

Premature neonates in neonatal intensive care units (NICUs) face frequent exposure to noxious stimuli, leading to acute, prolonged, or chronic pain. Despite its impact, prolonged pain remains poorly defined and managed due to inconsistent terminology in the literature. This concept analysis seeks to establish a clear theoretical definition of prolonged pain in premature neonates. Using Smith and Mörelius’ [1] principle-based methodology, 73 articles were systematically reviewed across eleven multidisciplinary health databases. Deductive analysis based on Anand’s[2] theoretical pain framework, combined with inductive analysis of pragmatic, linguistic, and logical principles, identified key elements to define prolonged pain. This definition will improve the identification, evaluation, and treatment of prolonged pain in hospitalized premature infants while enhancing both the scientific and clinical understanding of their pain experiences.

## Introduction

Premature neonates account for approximately 10 % of births globally, representing 13.4 million infants worldwide in 2020 [3]. Many premature neonates will require hospitalisation in a neonatal intensive care unit (NICU), exposing them to noxious stimuli. Premature neonates can feel pain starting at 24 weeks of gestation [4] and can experience pain more intensely than adults due to the immaturity of their nervous system [5]. A recent study showed that premature neonates can experience up to 25 acute painful procedures daily and up to 41 hours of cumulative chronic pain exposure per day due to concomitant medical procedures during the first 28 days of hospitalisation in the NICU [6]. The recommendations of the American Academy of Pediatrics (AAP) [7] and the Canadian Paediatric Society (CPS) [8] are to adequately evaluate and alleviate pain in order to prevent long term consequences on the neurodevelopment of premature neonates. Despite the recommendations from the AAP and the CPS, an observational study in 18 European countries and 243 NICUs showed that only 10 % of premature neonates were evaluated for prolonged pain daily and approximately two thirds of these premature neonates were not evaluated for prolong pain at all during their hospitalisation [9]. Currently in neonatology, a lack of consensus on a definition, as well as ambiguous terminology in the scientific literature may account for the absence of prolonged pain assessment in premature neonates by health care professionals and may further hinders the ability of these professionals to adequately evaluate and manage prolonged pain [2,10]. Due to the ambiguous terminology [10], “chronic pain” is often used interchangeably to describe the prolonged pain experience of premature neonates. However, the definition of chronic pain outlined by the International Association for the Study of Pain (IASP) [11] is pain that persists beyond three months. This definition clearly excludes premature neonates less than three months of life, emphasizing that a conceptual understanding of a prolonged pain state, which may lead to chronic pain, is necessary.

Researchers have attempted to define chronic pain in the premature neonate, but more work is needed in order to distinguish transitional states of pain such as prolonged pain or persistent pain [10,12-13]. Anand [2] presents an initial starting point for defining non acute pain states such as persistent pain, prolonged pain and chronic pain. In this framework, Anand [2] defines neonatal prolonged pain having a rapid but gradual onset, lasting one to 24 hours, with a sharp and diffusely localised character and the possible presence of hyperalgesia. This initial definition is important to consider, but Anand [2] states that the temporal features of these pain state were arbitrarily defined at best, thereby illustrating the crucial need to further analyse and advance our comprehension of the concept of prolonged pain in premature neonates. Thus, the aim of this concept analysis is to clarify and define the concept of prolonged pain in premature neonates in order to operationalize a definition and, as a result, standardize the terminology used in the scientific literature and the clinical setting.

## Methods

This systematic knowledge synthesis uses the principle-based approach to concept analysis by exploring the four philosophical principles of epistemology, pragmatic, linguistic, and logic outlined by the three-phased methodology of Smith and Mörelius [1]. The four philosophical principles are further discussed through conceptual components such as preconditions, characteristics and outcomes [1]. This approach, initially developed by Morse et al. [14] and further operationalized by Hupcey and Penrod [15], was favored over a more classical approach, such as the Wilson method [16], due to its robustness and thoroughness in evaluating the state of the scientific literature [17], its description of a systematic three-phased approach to developing a theoretical definition, and its quality evaluation of the included articles. The concept analysis protocol is registered on Open Science Framework [18]. Smith and Mörelius [1] describe four stages for each of the main three phases of the concept analysis such as preparation, analysis and results (see **Table 1**.). However, certain elements of the methodology such as pilot testing the article screening were not detailed in this method, therefore the Cochrane guidelines for systematic reviews [19] were used to guide these steps and the recommendations were integrated into the Smith and Mörelius [1] concept analysis methodology (see **Table 1**.). This systematic concept analysis is reported based on the PRISMA systematic review guidelines [20].

**Table 1.** An Adapted Phased Approach to Conducting a Principle-Based Concept Analysis.

| Phase | Stage 1 | Stage 2 | Stage 3 | Stage 4 |
| --- | --- | --- | --- | --- |
| **1. Preparation** | Determine the concept of interest (gaps in the literature or existence of a previously conducted concept analysis) | Protocol development.<br>Registration on Open Science Framework (OSF)[18]. | Development of research strategy equation.<br>Systematic literature search. | As per Cochrane recommendations [19]:<br>1.Pilot test 10 articles for screening to clarify inclusion/exclusion criteria.<br>2.Screen articles by title/abstract and full text by two independent reviewers. |
| **2. Analysis** | As per Cochrane recommendations [19]:<br>1.Pilot test 5 articles with data extraction table.<br>2.Data extraction by two independent reviewers and consensus obtained by a third independent reviewer.<br>Initial notetaking for each of the four principles. | Pilot test 5 articles for the relevance assessment tool. | Perform quality criteria assessment by two independent reviewers. Conflicts resolved by group discussion. | Integrate data following a recommended qualitative inductive analysis. |
| **3. Results** | Presentation of the quality criteria assessment findings for each article. | Summary of the conclusions for the four principles. | Explore the conceptual components (preconditions, characteristics, and outcomes). | Formulate a theoretical definition. |

*Note.* This table was modified from Smith and Mörelus (2021, p.5) [1].

### Eligibility Criteria

Articles discussing prolonged pain in premature neonates that are hospitalised in the NICU were included in this concept analysis. Prematurity is defined by the World Health Organisation [21] as neonates born at less than 37 completed weeks of gestational age. Articles discussing both term and premature neonates were included, and special attention was made to extract data only pertaining to premature neonates. Articles discussing “ongoing”, “persistent”, ‘continuous”, and “repetitive pain” were included if the studied pain did not persist beyond three months. Articles discussing only pain persisting beyond three months in premature neonates were excluded as they relate to the definition of chronic pain outlined by the IASP [11] and therefore do not contribute to defining prolonged pain. Articles in English or French were included and were any of the following types: quantitative, qualitative or mixed-methods studies, knowledge syntheses, opinion articles or thesis dissertations.

### Information Sources

This concept analysis elaborates on the findings of a previous scoping review of neonatal prolonged pain conducted by the main author [22] and uses the same inclusion and exclusion criteria. Therefore, 26 articles defining neonatal prolonged pain identified in this scoping review, searched from inception to November 20, 2022, in MEDLINE, PubMed, CINAHL, Web of Science and the grey literature (Grey Lit. Org, Grey Source Index), were included in this concept analysis. An updated search was conducted on March 24^th^, 2024, in these databases, using specific field codes to identify articles entered into the database [23] since the last search on November 20^th^, 2022. An additional search was conducted from inception to March 24^th^, 2024, in PsychInfo, EMBASE, ProQuest, BMJ best practice, JBI EBP database and EBM reviews/Cochrane. Manual searches of the reference lists of articles written by key authors researching neonatal prolonged pain such as Anand [2], Ilhan [10,24-26], Pillai Riddell [12], and van Ganzewinkel [13,27], were conducted to identify any articles not obtained through the searches.

### Search Strategy

The search strategy launched in CINAHL with MH Exact Subject heading and keywords such as “premature neonate” and “prolonged pain” can be found in the registered protocol [18]. The search strategy was developed with consultation with the nursing sciences librarian from the Faculty of Nursing at the Université de Montreal. The search strategy included terms such as “infant” and “newborn” because relevant articles discussing premature neonates are often indexed under these terms in databases based on the author keywords. The search strategy was then launched in the remaining databases and was adjusted to meet the requirements of each database. No limits of language or year of publication were applied to the searches.

### Selection process

All articles identified by the search strategy were uploaded into the Covidence systematic review software (Veritas Health Innovation, Melbourne, Australia, www.covidence.org), where duplicates were removed. The articles’ selection followed a two-step process by two independent reviewers in duplicate, where the title and abstracts were screened for eligibility followed by the review of full texts and reasons for exclusion were reported. As recommended by Cochrane, an initial sample of 10 articles was piloted (which included articles that were a definite No, a definite Yes and a Maybe) in order to clarify the eligibility criteria, to train the reviewers and to ensure that the criteria were applied consistently among the reviewers [19]. Any disagreements were resolved by discussion or a third independent reviewer. The results of the selection process is shown by a PRISMA flow diagram (see **Figure 1**.).

**Figure 1.**
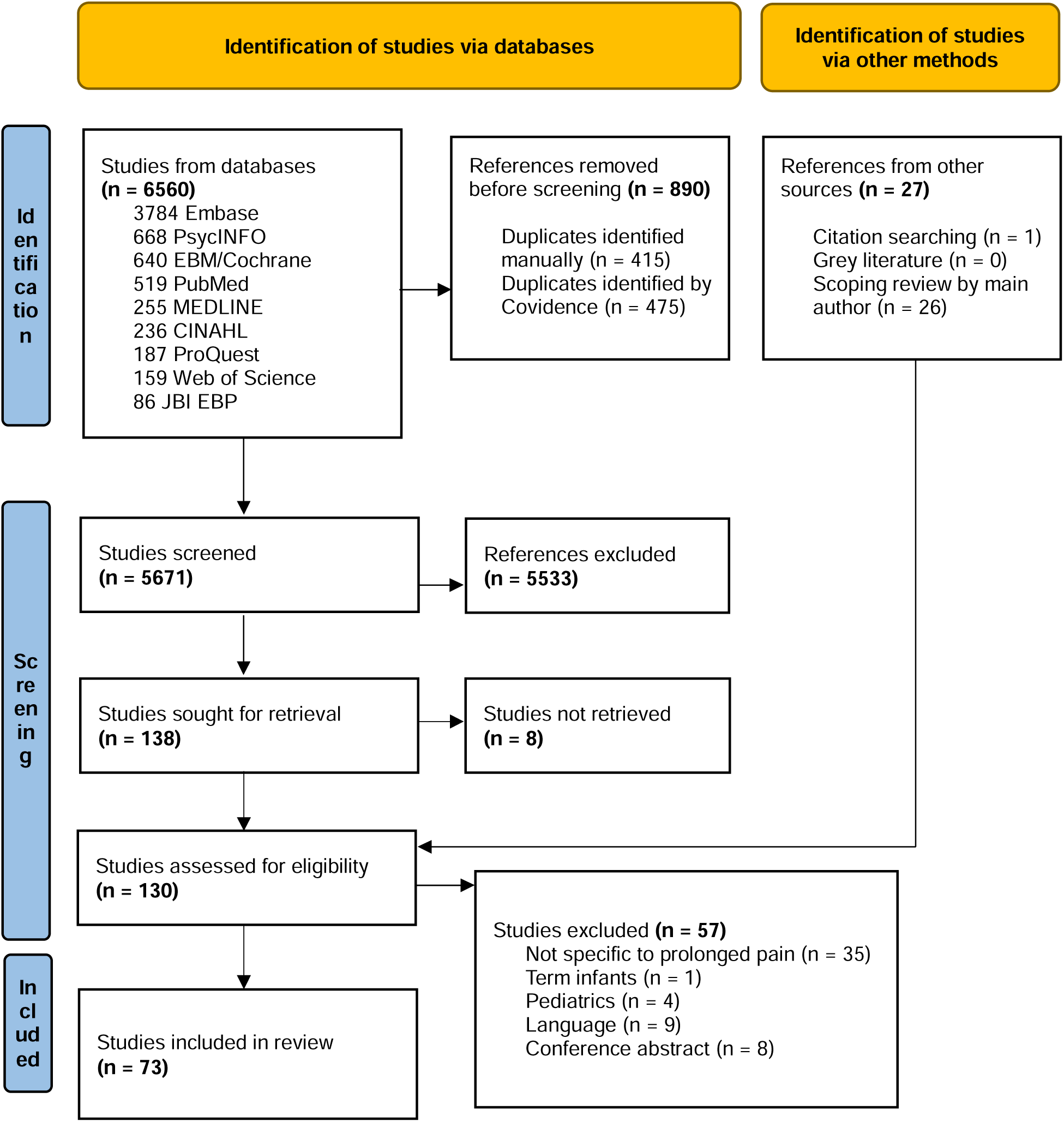
PRISMA flow diagram of the database search conducted on March 24^th^, 2024, reported as per the PRISMA guidelines [20].

### Data extraction and data items

Data was extracted by two independent reviewers in duplicate in a table developed by the main author (ABP) in Covidence. The extraction table underwent pilot testing process with a sample of 7 articles (representing 10 % of the articles included) by two independent reviewers. A sample of 10 % was randomly selected as no quantity was specified in the concept analysis methodology[1] and a recent methodological review of systematic review data extraction forms was also inconclusive in quantifying the number of extraction forms to pilot [28]. Data extracted included author(s), country and year of publication, study design, population, information regarding the four principles of Smith and Mörelius [1], namely epistemology pragmatism, linguistics, and logic as well as the conceptual components. The principles and conceptual components are defined in **Table 2**.

**Table 2.** Principles and Conceptual Components as described by Smith and Mörelius [1].

| ***Principles*** | **Definition** |
| --- | --- |
| Epistemology | Epistemological data includes any definitions, characteristics, or time criterion of prolonged pain. |
| Pragmatic | Pragmatic information includes any tools or methods to measure prolonged pain in premature neonates. |
| Linguistic | Linguistic refers to terms used to describe prolonged pain. |
| Logic | Logic refers to any theories or conceptual boundaries of the concept of neonatal prolonged pain. |
| ***Conceptual Components*** |  |
| Precondition | Phenomena that precedes or influences neonatal prolonged pain. |
| Characteristic | Frequent words or expressions used to describe the experience of neonatal prolonged pain. |
| Outcomes | The consequences that follow the occurrence of neonatal prolonged pain. |

### Quality appraisal

Quality appraisal was conducted using the quality criteria tool developed by Smith and Mörelius [1] to evaluate the relevancy of the articles in providing information regarding the four principles of concept analysis. As suggested by Smith and Mörelius [1], the quality criteria tool was piloted with five articles, and this was carried out by the entire research team independently and in duplicate. No articles were excluded based on the quality criteria tool score, but these scores were considered in the discussion of the results and are tabulated in Table 3.

**Table 3.**
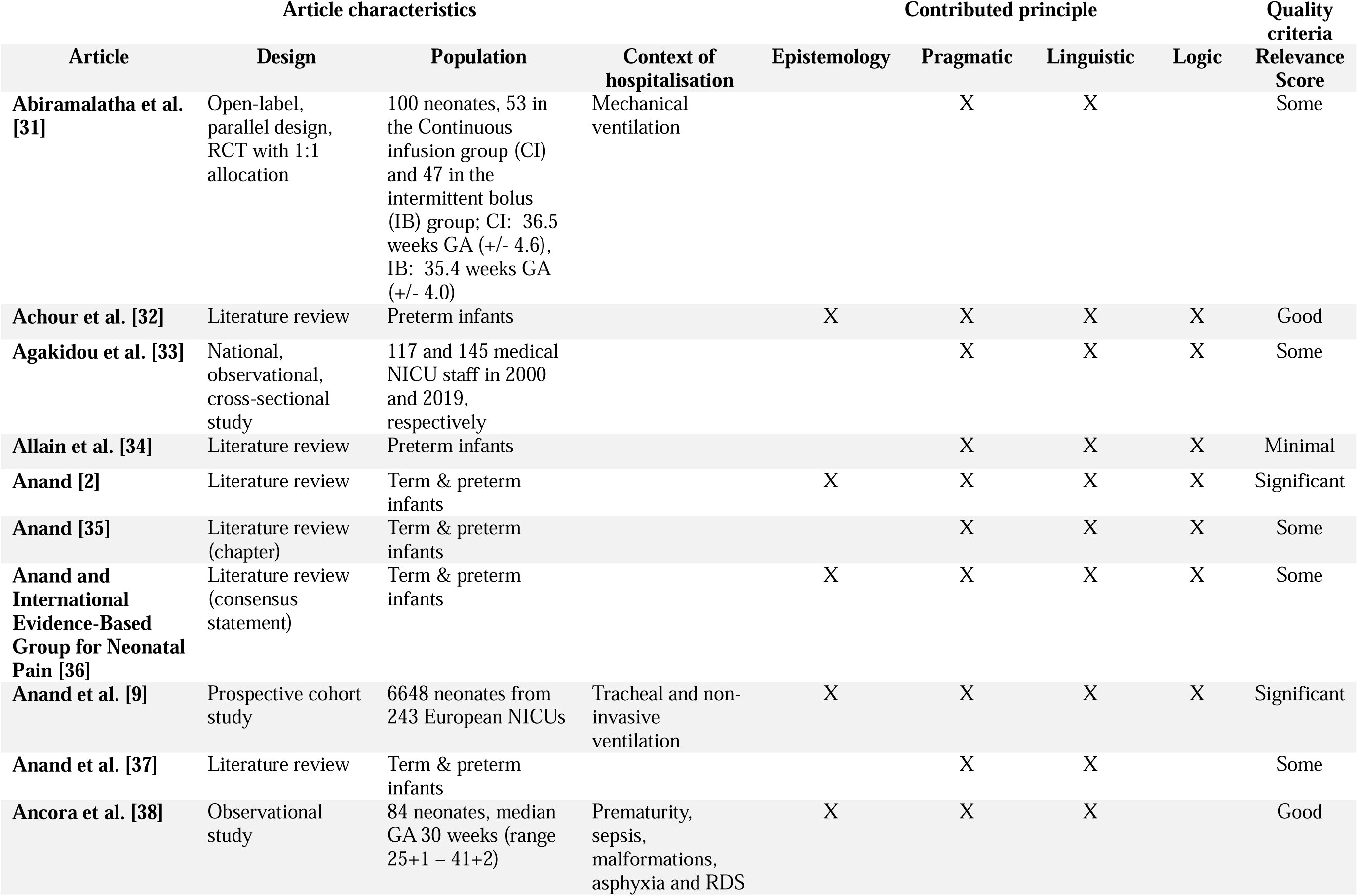
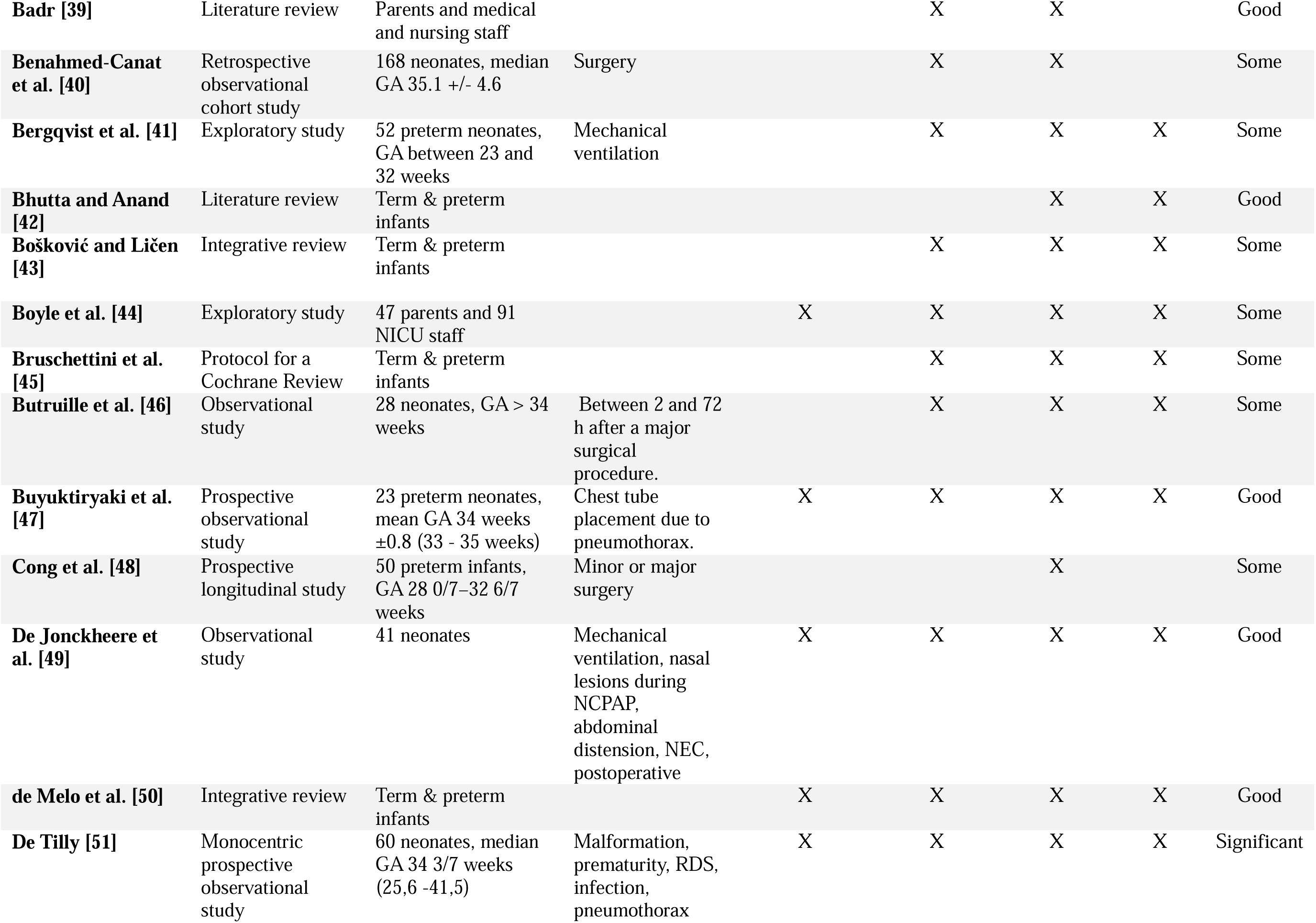
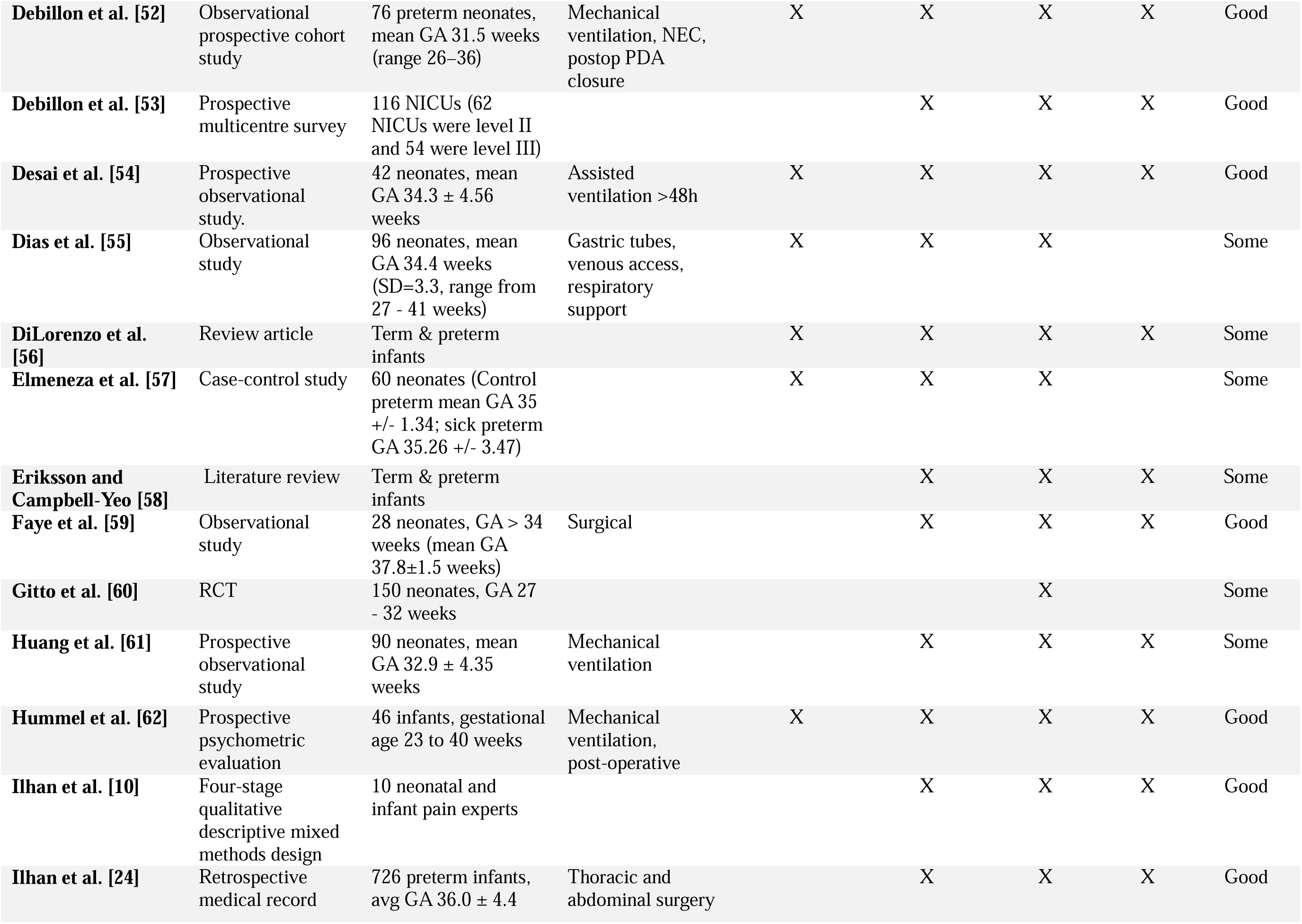
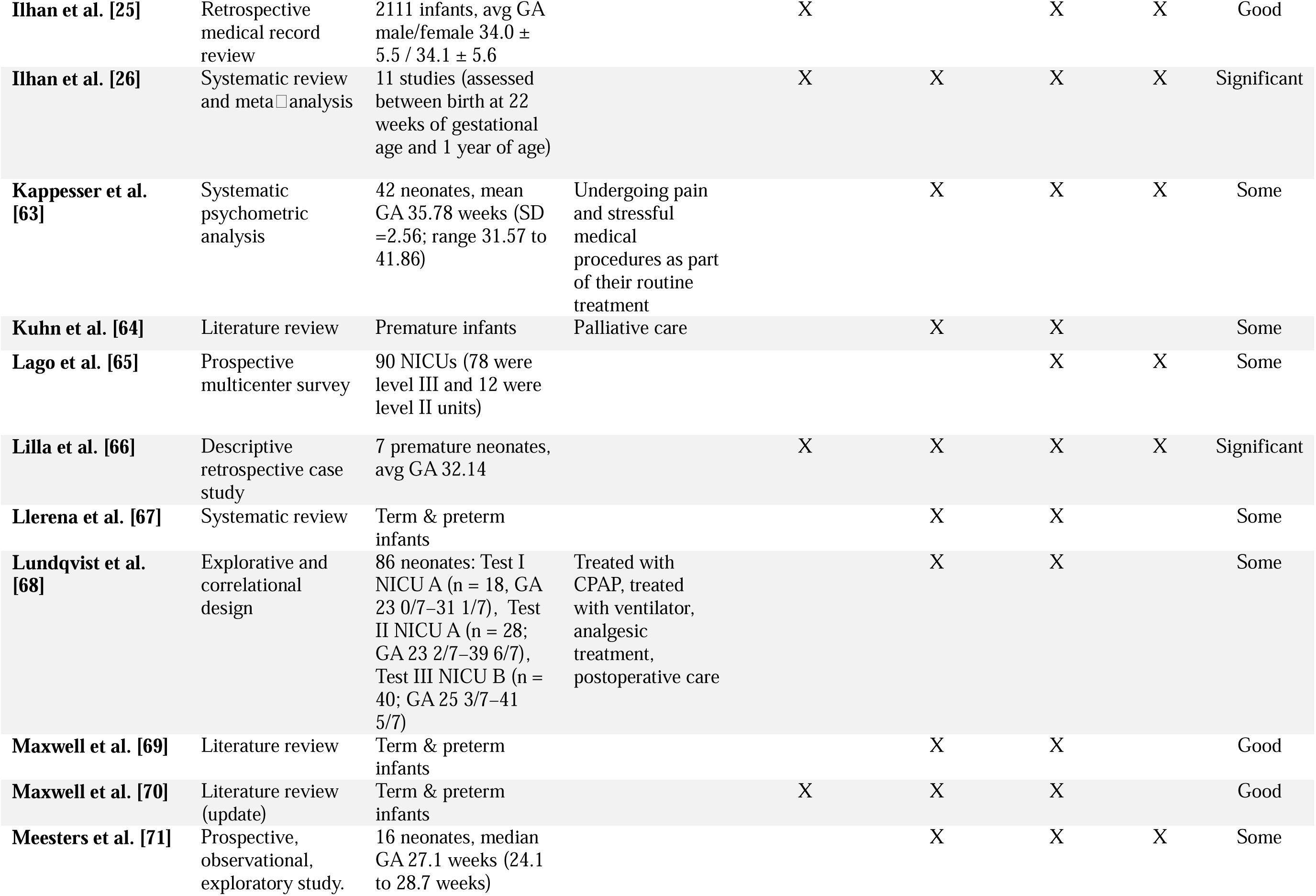
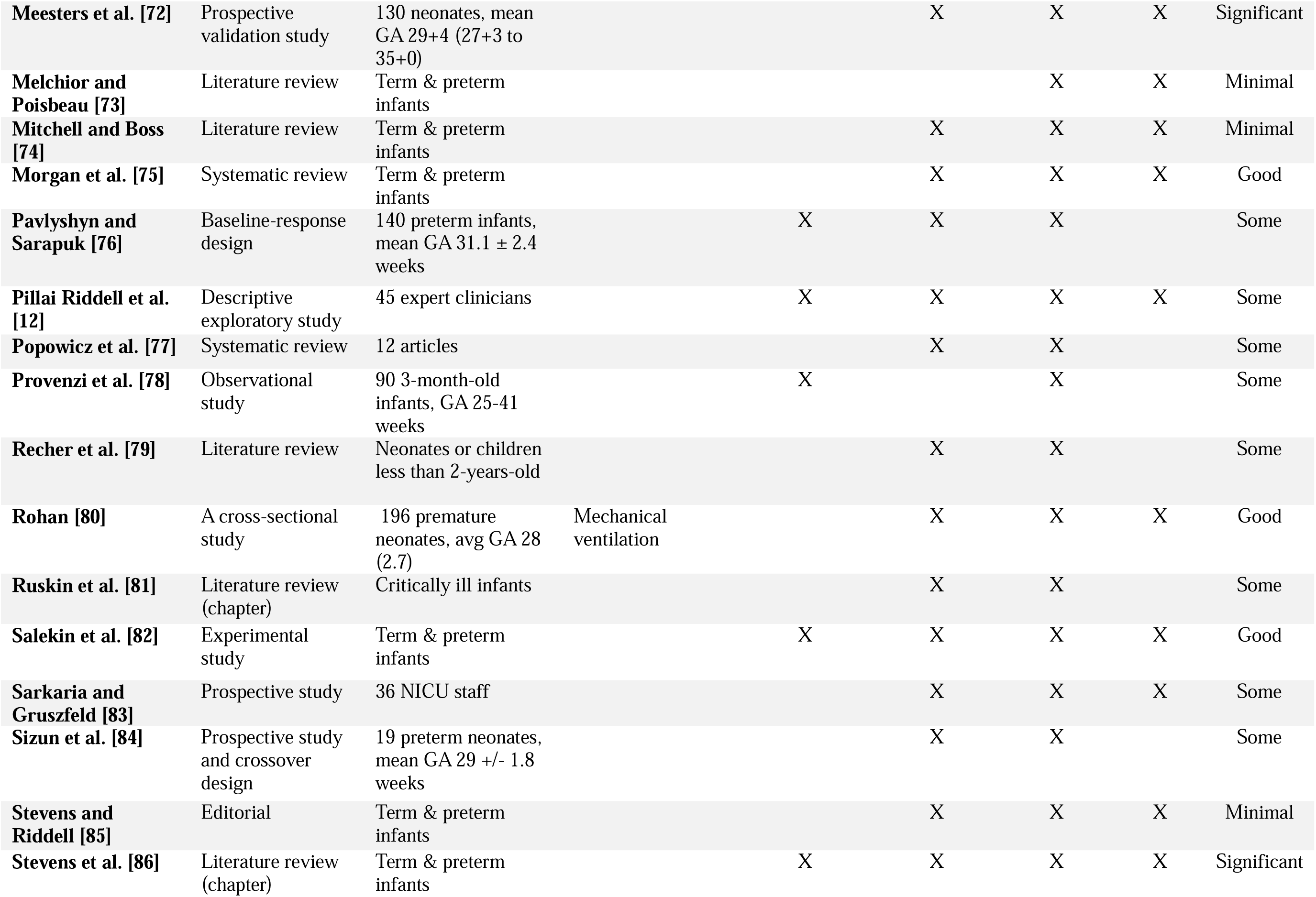
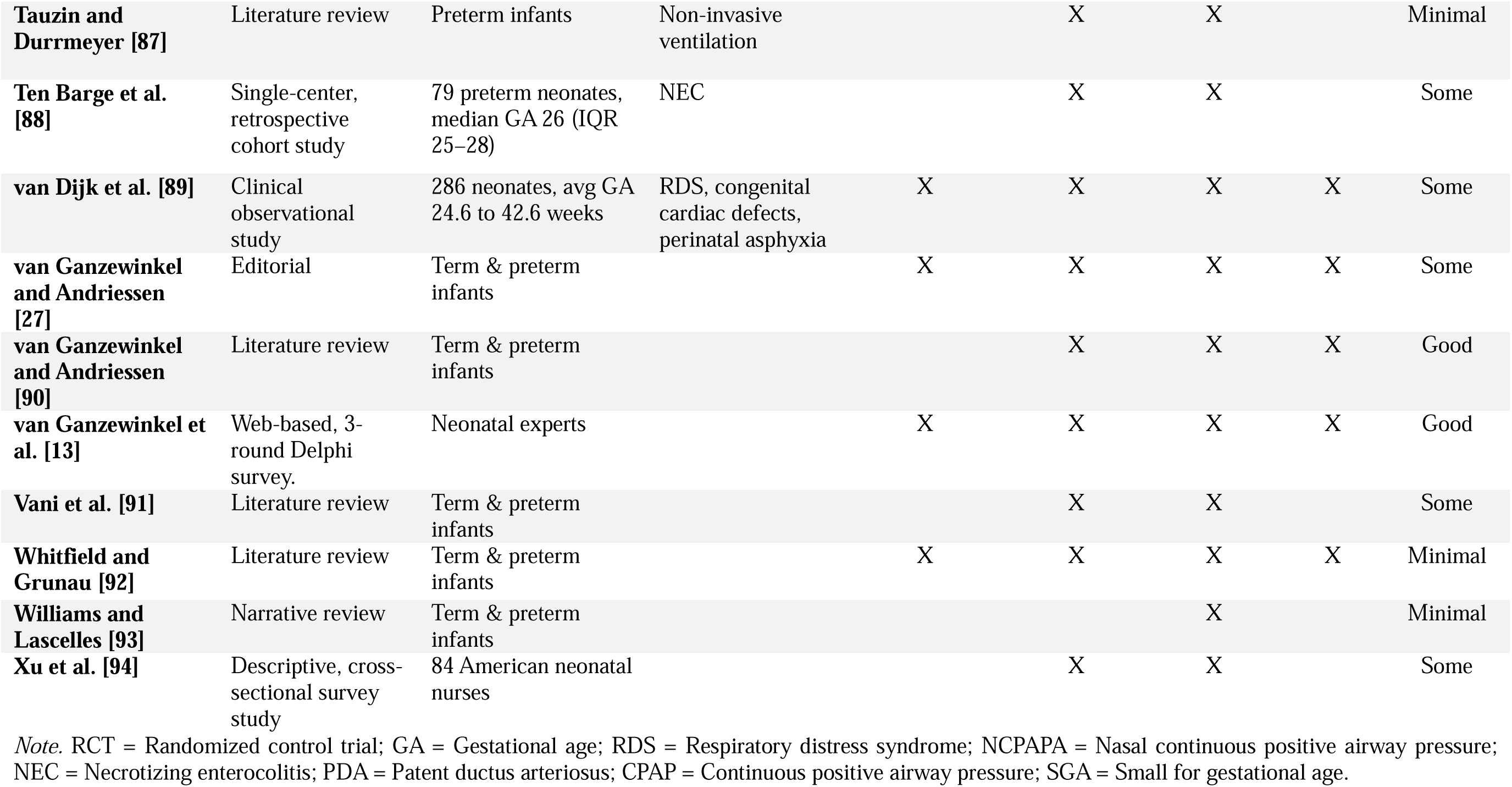
Descriptive table of included articles.

### Data analysis

Initial note taking was done based on the four principles of epistemology, pragmatic, linguistic, and logic. More specifically, the information pertaining to the epistemological principle was analysed based on the deductive content analysis approach described by Hsieh and Shannon [29]. The goal of deductive content analysis is to validate and extend conceptually a theoretical framework [29]. Based on the previous works defining non acute pain in premature neonates, this concept analysis contributes to further validate and extend the theoretical framework put forth by Anand [2]. Anand [2] sought to differentiate various types of neonatal pain into five pain states (acute episodic, acute recurrent, prolonged, persistent, and chronic) with distinct categories such as onset, duration, character, primary hyperalgesia, secondary hyperalgesia, allodynia, behavioural phenotype, and physiological phenotype. Following the deductive content analysis method described by Hsieh and Shannon [29], the information was coded based on predetermined categories aforementioned and outlined in Anand’s [2] framework of neonatal pain. Next, the information pertaining to the pragmatic, linguistic and logic principles were analysed with an inductive content analysis approach, also described by Hsieh and Shannon [29]. This inductive approach is more relevant for these principles, as previous research on these topics is limited and an inductive category development approach allows the categories to flow from the data itself [29]. Finally, the frequency of recurring information was calculated for each category, for both the deductive and inductive approaches, to present in ranking order, the most important information pertaining to the four principles of concept analysis, as determined by the different pain researchers in their articles.

From a realist ontological and a moderate objectivist epistemological perspective, validating previous work that attempts to define neonatal prolonged pain and thereby elaborating on current knowledge, will allow for the theoretical evolution and advancement of the definition of neonatal prolonged pain. This is also a pragmatic approach to concept analysis in that it contributes to the usefulness of Anand’s [2] neonatal pain framework. The computation of frequencies of occurrence confers the benefits of revealing a consensus amongst the scientific literature and offering objective results. Also, it aligns with a pragmatic philosophical perspective in that there is no absolute truth but rather a consensus in constant evolution of what is the best and most useful information [30] for defining neonatal prolonged pain.

## Results

The systematic search and selection of articles resulted in 73 articles being included in this concept analysis, which are described in **Table 3**. The concept of prolonged pain is a recent concept in neonatology. More specifically, some articles were published before 2010 (n = 20/73; 27.4%), more than half of the articles were published between 2010 and 2020 (n = 39/73; 53.4%) and a few articles were published after 2020 (n = 14/73; 19.2%). Most of the articles were knowledge syntheses (n = 29/73; 39.7%), with few systematic reviews (n = 4/73; 5.5 %). The remaining articles were predominantly observational studies (n = 26/73; 35.6%), such as validation studies for pain tools or retrospective medical chart reviews, while very few experimental studies explored the concept of neonatal prolonged pain (n =5/73;6.8 %), where only two (n = 2/73; 2.7 %) were randomized control trials. The articles included were conducted or published in the United States of America (n = 20/73; 27.4 %), France (n = 12/73; 16.4%), The Netherlands (n = 7/73; 9.6%), Canada (n = 6/73; 8.2%), and Australia (n = 4/73; 5.5%). The quality criteria tool scores are shown in **Table 3**. and a detailed quality assessment of each article against the four principles can be found in the supplemental files (S1). Only seven articles (n = 7/73; 9.6%) provided significant information for the understanding of the concept of neonatal prolonged pain, whereas some articles provided good information (n = 23/73; 31.5%), half of the articles provided some information (n = 36/73; 49.3%) and some provided minimal information (n= 7/73; 9.6%) for the understanding of our understanding concept of neonatal prolonged pain.

## Principles

### Epistemology

The principle of epistemology was the least discussed of the four principles, with more than a third of the included articles conferring information contributing directly to the defining information of prolonged pain in premature neonates (n=29/73; 39.7%). The results are presented following Anand’s [2] pain framework which is comprised of the following categories: onset and duration, character, primary and secondary hyperalgesia, allodynia, behavioural phenotype and physiological phenotype.

## Onset & Duration

The temporal features of duration (n = 16/29; 55.2%), and onset were the most discussed elements in articles (n = 10/29; 34.5%). Pertaining to the onset of prolonged pain, most articles mentioned that prolonged pain resulted from a clear stimulus, such as medical, surgical or therapeutic procedures (n = 6/10; 60 %) and only three articles specified that prolonged pain was not dependent on a noxious stimulus (n = 3/10; 30%). In terms of duration, most articles stated that prolonged pain lasted a longer time (n = 7/16; 43.8%), had a definable beginning and end point (n = 4/16; 25%), lasted several hours or days (n = 5/16; 31.3%) and needed a longer healing time than acute pain (n = 3/16; 18.8%).

## Character

The character of prolonged pain was discussed by only three articles that addressed epistemological elements of a definition of prolonged pain (n = 3/29; 10.3%). The character of prolonged pain was stated in the framework by Anand[2] and described as being sharp and diffusely localised. Whereas two articles stated that prolonged pain fluctuates in character (n = 2/3; 66.7%), with one article specifying that prolonged pain repeats with decreasing intensity (n = 1/3; 33.3%).

## Primary & Secondary Hyperalgesia

Hyperalgesia, or increased pain to a stimulus that is normally painful, was discussed in two articles (n = 2/29; 6.9%). Primary hyperalgesia, or pain localised to area of injury, was deemed present in the context of prolonged pain in both articles. Secondary hyperalgesia, pain diffused to remote areas away from the site of injury, was estimated to be present or mildly present by both articles and possible prolonged by one article.

## Allodynia

Allodynia, pain from a stimulus that normally does not invoke pain, was discussed in two articles (n= 2/29; 6.9%). However, both articles were divergent in their findings, with one article stating allodynia was probably absent and the other mentioning that it was present and mild.

## Behavioural phenotype

Behavioural phenotypes were discussed in seven articles (n = 7/29; 24.1%). Behavioural phenotypes in premature infants may refer to consistent patterns of behaviour over time. Two articles stated it was important to observe behavioural activity in prolonged pain (n = 2/7; 28.6%). Two articles described the behavioural phenotype of prolonged pain as strongly reactive on stimulation or hypersensitive (n = 2/7; 28.6%). On the other hand, two articles stated that the behavioural phenotype has a dwindling reactivity (n = 2/7; 28.6%), and one article specified that premature neonates would adapt to prolonged pain from a behavioural standpoint (n = 1/7; 14.3%).

## Physiological phenotype

Physiological phenotypes of prolonged pain were discussed in eight articles (n = 8/29; 27.6%). Most of the articles stated that premature neonates may adapt to prolonged pain from a physiological standpoint as presented by an altered or permanent shift in basal autonomic arousal (n = 6/8; 75%). One study further specified that a lower variability in heart could be observed (n = 1/8; 12.5%) and one study mentioned that the HPA axis reactivity may be altered in premature neonates with prolonged pain.

### Pragmatic

Most of the included articles discussed pragmatic information (n = 65/73; 89%). Inductive analysis of pragmatic information revealed five categories: pain evaluation tools, behavioural indicators, physiological indicators, contextual indicators and pain assessment of prolonged pain.

## Pain evaluation tools

Fifty articles stated various pain evaluation tools that were validated or used to measure prolonged pain (n = 50/65; 76.9%). The following pain evaluation tools were frequently cited: Échelle de Douleur et Inconfort du Nouveau-né (EDIN) (n = 30/50; 60%), the Neonatal Pain and Agitation Sedation Scale (N-PASS; n = 26/50; 52%), the COMFORTneo scale (n = 15/50; 30%), the NIPE index or hear rate variability (HRV; 7/50; 14%), the COMFORT scale (n = 6/50; 12%), the Premature Infant Pain Profile (PIPP; n = 4/50; 8%) and the Pain Assessment Tool (PAT; n = 3/50; 6%).

## Behavioural indicators

Behavioural indicators were discussed in sixteen articles (n = 16/65; 23.1%). Facial expressions were most identified as being behavioural indicators of prolonged pain (n = 7/16; 43.8%). Other important behavioural indicators included high activity levels or body movements (n = 5/16; 31.2%), facial amimia, or reduced facial expressions (n = 3/16; 18.8%) and cries of longer duration (n = 3/16; 18.8%). On the other hand, crying was noted to not be a relevant parameter for prolonged pain as it is not energy efficient (n = 2/16; 12.5%). Interestingly, some articles stated that clear and specific signs of prolonged pain remain undefined (n = 4/16; 25%). Of note, certain articles (n = 2/16; 12.5%) specified other behavioural indicators such as the position of extremities such as finger clenching or splaying, muscle tone, poor response to handling, poor ventilator synchronicity, agitation or irritability, consolability, sociability, quality of sleep and low-grade distress due to poor energy reserves. Each of the following behavioral indicators was discussed in a single study, though across different articles (n = 1/16; 6.3%): decreased response to further pain events, withdrawal, lower threshold for pain and passivity with no limb movement due to energy depletion from the underlying disease.

## Physiological indicators

Physiological indicators were discussed in seventeen articles (n = 17/65; 26.2%). Almost half of the articles discussing physiological indicators agreed that these indicators are not present or appropriate for prolonged pain (n = 8/17; 47.1%). A considerable number of articles mentioned that heart rate variability, with tools such as the NIPE index, would be diminished (n = 7/17; 41.2%). More specific physiological indicators were briefly mentioned by certain articles such as proto-oncogenes such as c-fos (n = 1/17; 5.9%) or levels of neuropeptide substance P (n = 1/17; 5.9%) as markers for pain.

## Contextual indicators

Contextual indicators were discussed in thirty-five articles (n = 35/65; 53.1%). Many key contexts of hospitalisation were identified as contextual indicators of prolonged pain such as mechanical ventilation (n = 20/35; 57.1%), postoperative period (n = 13/35; 37.1%), necrotizing enterocolitis (n = 10/35; 28.6%), repetitive iatrogenic acute procedures (n = 7/35; 20%), inflammatory conditions (n = 4/35; 11.4%), NICU hospitalisation (n = 3/35; 8.6%) and chest tube insertion and/or drainage (n = 2/35; 5.7%). Other interesting contexts of hospitalisation identified but discussed briefly in singular articles (n = 1/35; 2.9%) for example, osteogenesis imperfecta, epidermolysis bullosa, abdominal distention during enteral nutrition, nasal lesions during nasal CPAP, acute osteomyelitis, intracranial hypertension, birth trauma, poorly controlled acute pain, CPAP or high flow and nerve damage. Other contextual indicators included gestational age (n = 3/35; 8.6%), severity of illness (n = 1/35; 2.9%), as well as environmental factors such as noise (n = 1/35; 2.9%).

## Pain assessment

Pragmatic information pertaining to pain assessment was identified in twenty-one articles (n = 21/65; 32.3%). It was deemed important to provide continuous assessment of prolonged pain rather than periodic scoring (n = 5/21; 23.8%). It was also noteworthy that mechanical ventilation, sedation and severity of illness might mask pain responses (n = 3/21; 14.3%). Two other articles (n = 2/21; 9.5%) mentioned elements impacting pain assessment practices, such as the need to provide a multimodal assessment of prolonged pain, the importance of prolonged pain protocols for increasing evaluation and management, as well as the disassociation between behavioural and physiological pain responses. Further interesting elements of pain assessment briefly described by one article (n =1/21; 4.8%) included cutoff points for prolonged pain may not be valid for current pain assessment tools, nurses are key person for identifying pain, consolability requires nursing intervention, a one-size fits all approach to prolonged pain assessment is unsatisfactory and sex differences in pain responses exist.

### Linguistic

The inductive analysis of the linguistic principle was concerned with the consistency in use and meaning, as well as the fit of the concept of neonatal prolonged pain across a variety of contexts. Neonatal prolonged pain was consistently used across all included articles to describe a prolonged pain state in premature neonates hospitalised in the NICU in various medical contexts (see **Table 3**.). Variability was observed in the terminology used to describe the prolonged pain state of the premature neonate. Identified pain terms included “prolonged pain”, “repetitive pain”, “continuous pain”, “persistent pain”, “long term pain”, “non-acute pain”, “ongoing pain”, “acute continuous”, “acute prolonged”, “postoperative pain”, “iatrogenically prolonged pain”, “chronic pain”, “cumulative pain” and “prolonged procedural pain”. Most articles (n= 66/73; 90.4%) used prolonged pain to describe a non-acute pain state, thirteen articles (n = 13/73; 17.8%), were more specific and also referred to prolonged pain as “acute prolonged”, twelve articles used “ongoing pain” (n = 12/73; 16.4%), eleven articles used the term “persistent pain” (n = 11/73; 15.1%), six articles referred to “continuous pain” (n = 6/73; 8.2%) and seven articles used “chronic pain” and “prolonged pain” interchangeably (n = 7/73; 9.6%). Interestingly, more than half of the articles (n = 38/73; 52.1%) consistently used only one term to describe the prolonged pain state being studied or evaluated, whereas approximately a quarter of studies (n = 20/73; 27%) used two different terms to refer to prolonged pain and the remaining articles (n = 15/73; 20.5%) used three or more pain terms to describe a prolonged pain state. **Figure 2**. represents visually the variability in the terminology used by authors in the included studies.

**Figure 2.**
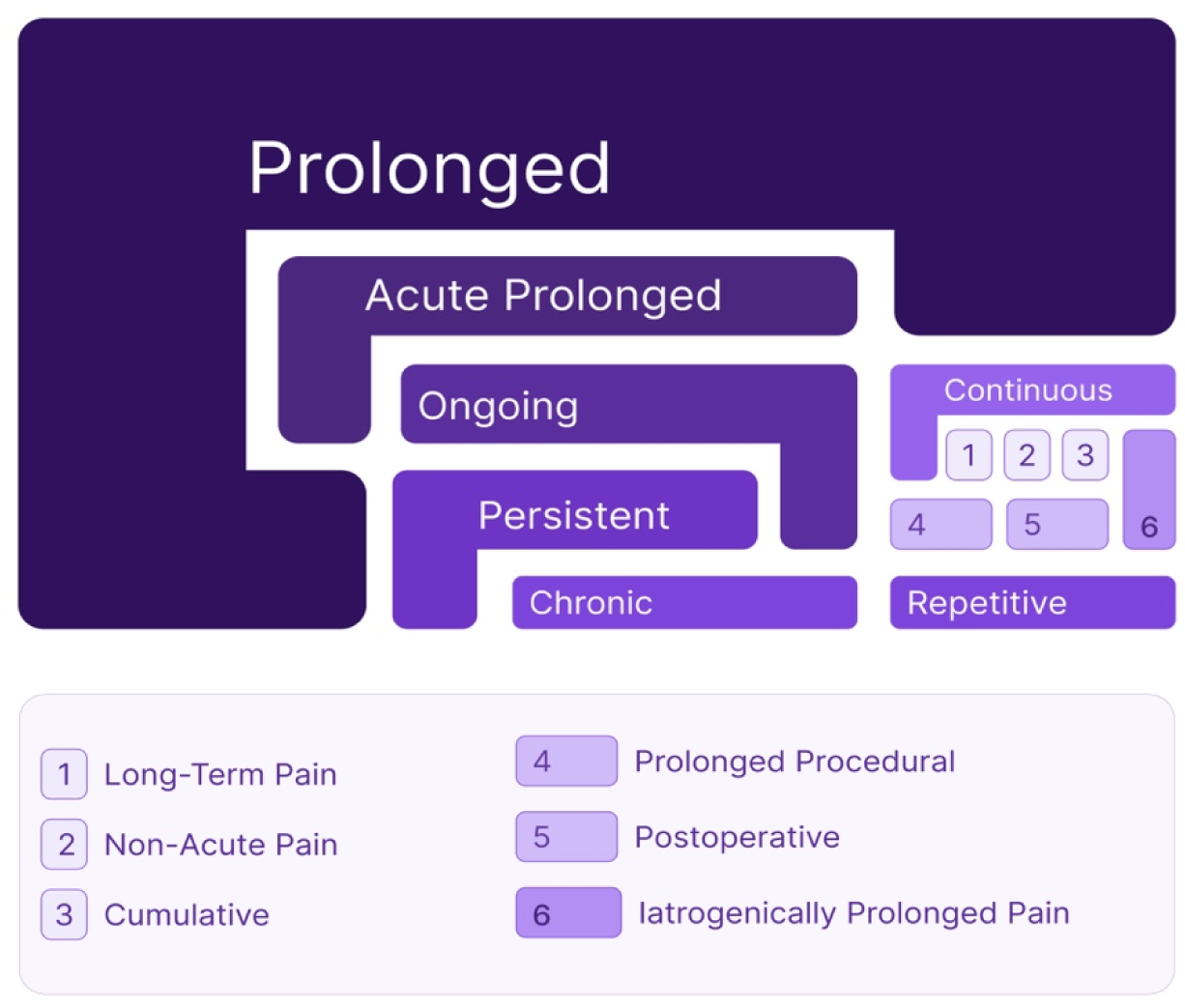
Weighted representation of the terminology used to describe prolonged pain in the 73 articles included in this concept analysis.

### Logic

An inductive analysis of the logic principle revealed two categories, specifically, theoretical frameworks (n = 8/48; 16.7%), and conceptual boundaries (n = 42/48; 87.5%) in more than half of the articles included in this concept analysis (n = 48/73; 65.8%). Theoretical frameworks discussed included the Windup theory in the presence of prolonged pain (n = 3/8; 37.5 %), the Synactive theory of development (n = 2/8; 25%) and the Gate control theory (n = 2/8;25%). Biological principles were also discussed such as brain plasticity and brain immaturity (n = 2/8; 25%), the complex interaction of biopsychosocial mechanisms (n = 2/8;25%), NMDA mediated excitotoxicity (n = 2/8; 25%), and the secretion of neuropeptide substance P (n = 1/8; 12.5%).

Conceptual boundaries included articles where prolonged pain was deemed conceptually and theoretically different than other pain states. Close to half of the articles distinguished acute pain from prolonged pain (n = 17/42; 40.5%). Six articles categorized pain into three types: acute procedural, acute prolonged and chronic pain (n = 6/42; 14.3%). Six articles used Anand’s pain framework which distinguished acute episodic, acute recurrent, persistent, prolonged and chronic (n = 6/42; 14.3%). Four articles differentiated various non-acute pain states such as persistent pain, prolonged pain, and chronic pain (n = 4/42; 9.5%). Four articles distinguished three types of pain such as acute/procedural, prolonged and chronic pain and another four articles distinguished postoperative pain from prolonged pain (n = 4/42; 9.5%). One article described prolonged pain as being a transitional state of pain between acute and chronic pain (n = 1/42; 2.4%).

## Discussion

The concept of prolonged pain in premature neonates is relatively novel in the field of neonatology, especially when compared to the longer-standing recognition of acute pain. Most of the articles included in this review were published within the last 15 years, reflecting the emerging interest in this specific aspect of neonatal pain. While prolonged pain in premature neonates was formally and collectively recognized by the neonatal community as early as 2001 in the Consensus statement for the prevention and management of pain in the newborn by Anand and the International Evidenced-Based Group for Neonatal Pain [36], a definition of prolonged pain has not achieved a consensus among researchers and clinicians [2,10,13]. This concept analysis identified consensus elements pertaining to four principles from a variety of sources, mainly literature reviews and observational studies published predominantly in North America and Europe. These consensus elements allowed for a clearer understanding of the conceptual components of the concept of prolonged pain, more specifically the preconditions, the characteristics and the outcomes [1]. As a results, the conceptual components identified lead to the formulation of a theoretical definition of prolonged pain in premature neonates presented in the same format used by the IASP.

### Preconditions

Premature neonates are born during a crucial period of neurodevelopmental plasticity [95] and the preterm brain is vulnerable to a heightened pain experience in part due to peripheral sensitization, immature descending inhibitory pathways and overlapping receptive fields [35,93]. In addition to the cerebral immaturity, exposure to repetitive iatrogenic procedures causes physiological disruptions that may be preconditional to prolonged pain through channels such as NMDA mediated excitotoxicity [42,74,96], windup phenomenon [35,74,92-93] as well as a lack of homeostasis amongst subsystems as described by Als’ [97] Synactive Theory of Development. The pragmatic principle highlighted that the context of hospitalisation may be an important indicator of prolonged pain. Contexts such as mechanical ventilation, the postoperative period, NEC and other inflammatory conditions may be preconditions to experiencing prolonged pain; findings that are consistent with a recent scoping review on prolonged pain in premature neonates [22].

### Characteristics

Through deductive analysis of the epistemology principle using Anand’s [2] framework, characteristics of prolonged pain were identified. Temporal aspects of prolonged pain, such as its duration and onset, were determined as key characteristics. This is supported by the fact that more than half of the articles defining prolonged pain included information clarifying its time-related criteria. Although Anand [2] acknowledges that the durations assigned to prolonged, persistent, and chronic pain in his neonatal pain framework are arbitrary at best, authors discussing the temporal features of prolonged pain in articles included in this concept analysis agreed that it typically lasts from several hours to several days, with an extended healing time beyond what is considered normal or expected given the context of hospitalisation. Furthermore, more than half of the authors discussing the onset of prolonged pain agreed that it originated from a clear stimulus such as surgical, diagnostic or therapeutic procedures. This is consistent with the linguistic and logical principles, as many authors used terms and conceptual boundaries such as “acute prolonged”. While prolonged pain is not acute in its temporal features, it may be closely linked to a clear stimulus such as acute painful procedures or more largely a specific context.

The pragmatic principle highlighted consensus elements pertaining to the behavioural and physiological indicators. Many researchers agreed that maintaining physiological pain indicators such as tachycardia, hypertension, and tachypnea requires substantial energy, making them neither feasible nor appropriate markers for assessing prolonged pain in premature neonates [2,51,55,72,94]. Therefore, a strong behavioural phenotype is preferable for premature neonates experiencing prolonged pain [2]. Consensus behavioural indicators of prolonged pain include facial expressions, high activity levels, body movement (finger clenching, finger splaying, position of the extremities), muscle tone, response to handling, agitation and quality of sleep.

### Outcomes

The outcomes of prolonged pain in premature neonates highlighted by articles pertained specifically to neurodevelopmental consequences of untreated repetitive pain in early life. More immediate consequences of prolonged pain include hypersensitivity to otherwise non painful stimuli [93], negative impact on infant growth [9,66] , increased morbidity [32,74] and prolonged hospitalisation [9]. Long-term neurological consequences include impaired motor and cognitive development [13,66], impaired memory [13,94], altered pain processing and thresholds [2,57,60,93], attention deficit disorder [13,93], and a predisposition to the development of chronic pain as an adult [93].

### Theoretical Definition

Proposed here is a theoretical definition of prolonged pain in premature neonates, inspired by the framework suggested by Anand [2], considering the four principles and the conceptual components outlined by Smith and Mörelius [1] and representing the summarized findings from 73 articles on neonatal prolonged pain spanning the last 27 years in neonatal research (1997-2024).

##### Text Box. 1 Definition of prolonged pain in premature neonates

###### Prolonged Pain

Prolonged pain in premature neonates is characterized as a non-acute pain state originating from a clear stimulus. It endures for several hours or days, with fluctuating intensity, and persists beyond the expected healing period and depending on the context of hospitalisation.

###### Note

Prolonged pain can lead to both primary and secondary hyperalgesia. Additionally, it may cause a shift in basal autonomic arousal in premature neonates, reducing the reliability of physiological indicators for pain assessment. Consequently, behavioural and contextual indicators become crucial for accurate continuous pain evaluation and individualised management.

### Strengths and limitations

This is the first concept analysis in premature neonates addressing the pressing need to define the prolonged pain experience in premature neonates in order to improve its evaluation and management practices in the neonatal clinical settings. This concept analysis identified key components of prolonged pain—such as preconditions, characteristics, and outcomes—using a hybrid approach based on four philosophical principles: epistemology, pragmatism, logic, and linguistics. Through this method, a theoretical definition of prolonged pain was proposed that moves beyond simple time-based criteria or proximity to a specific painful event. Instead, it emphasizes the complex, individualized, and longitudinal nature of prolonged pain in premature neonates. The analysis also underscores the need for a multimodal and continuous approach to evaluating pain in this vulnerable population. A rigorous and systematic methodology was followed, delivering comprehensive information across the four principles. This innovative approach not only offered an objective framework to define an otherwise ambiguous concept but also facilitated a detailed examination of its conceptual components. Moreover, by analyzing the frequency of occurrence within the scientific literature, the study pragmatically illustrated the existing consensus on prolonged pain in premature neonates. In addition, all three phases of this concept analysis followed a systematic methodology outlined by Smith and Mörelius [1] as well as the recommendations by the Cochrane group for systematic reviews [19], ensuring reliability in this knowledge synthesis. Finally, this concept analysis builds on the work of key neonatal pain researchers, further validates the framework put forth by Anand and provides an operational definition of prolonged pain for clinical practice and scientific research.

Although the inductive analysis of the pragmatic, linguistic, and logical principles may be influenced subject to confirmation bias by the authors due to the ambiguous terminology used across the included articles, the computation of frequencies of occurrence revealed consensus elements amongst the scientific literature and presented objective results. Finally, following the quality criteria assessment of the 73 articles included by two independent reviewers, a significant number of articles (n = 52/73; 71.2%) presented some or minimal information for the advancement of our understanding concept of neonatal prolonged pain. This however presents a limitation of the research that currently exists in the scientific literature rather than a methodological limitation of this concept analysis.

### Implications for clinical practice

The proposed theoretical definition of prolonged pain in premature neonates contributes to standardising the language used in the scientific literature and the clinical setting amongst health care professionals. Given the temporal nature of prolonged pain lasting several hours to several days, a continuous[27] and multimodal (considering behavioural and contextual indicators) [82] pain evaluation is necessary. The International Evidence-Based Group for Neonatal Pain recommends performing a pain evaluation every 4 to 6 hours daily [36]. Consistent with the findings a recent scoping review on prolonged pain in premature neonates [22], the contexts of prolonged pain identified by this concept analysis such as mechanical ventilation, necrotizing enterocolitis, the post-operative period, and repetitive painful procedures should inform a nursing evaluation of prolonged pain minimally every 4 -6 hours with pain evaluation tools such as the EDIN, the N-PASS or the COMFORTneo given nurses’ strategic position in pain evaluation in this vulnerable population. These findings will also contribute to developing prolonged pain management protocols in the clinical setting and informing continuing education for health care professionals working in the NICU such as nursing and medical staff. Specifically, in relation to behavioural and physiological phenotypes and a proposed time criterion for a prolonged state of pain that is highly individualised to the premature neonate and their context of hospitalisation. This proposed theoretical definition will equally inform evaluation practices of prolonged pain in clinical practice and research. Due to the continuous, fluctuating and individualised nature of prolonged pain in premature neonates, innovative neurophysiological approaches for continuous pain measurement such as near-infrared spectroscopy, electroencephalography or functional MRI need to be explored in contexts of hospitalisation that increase the risk of exposure to prolonged pain. These neurophysiological approaches have been studied in acute pain in premature neonates [98], but further studies are required to evaluate the efficacy of these approaches in prolonged pain in premature neonates. Furthermore, this theoretical definition will guide researchers in studying the psychometric properties of pain evaluation tools and determining their sensitivity to measuring prolonged pain in a longitudinal manner, in addition to measuring the efficacy of pain-relieving interventions with validated tools in identified contexts of prolonged pain. Finally, this theoretical definition of prolonged pain can inform epidemiological studies and explorations of prolonged pain in understudied populations such as premature neonates with neurological impairments such as asphyxia, neonatal abstinence syndrome, intraventricular hemorrhage, and periventricular leukomalacia.

## Data Availability

All data produced in the present work are contained in the manuscript

## Author Contributions

ABP and MA conceptualised and designed the concept review. ABP developed the research equation and performed the literature search. ABP, BBP, AR and CC performed the screening of articles. ABP, BBP and AB extracted data and performed the quality criteria scoring. ABP analysed the data and wrote the manuscript. All authors edited, revised, and approved the final version of the manuscript.

## Acknowledgments

The authors would like the thank Assia Mourid, the librarian of the Faculty of Nursing at the Université de Montréal for her rigorous aid in developing the research equation. The authors would also like to thank Anne-Marie Chave, a master’s student at Concordia University for developing the graphical representation of the linguistic analysis.

---

| Section and Topic | Item # | Checklist item | Location where item is reported |
| --- | --- | --- | --- |
| **TITLE** |  |  |  |
| Title | 1 | Identify the report as a systematic review. | N/A |
| **ABSTRACT** |  |  |  |
| Abstract | 2 | See the PRISMA 2020 for Abstracts checklist. | N/A |
| **INTRODUCTION** |  |  |  |
| Rationale | 3 | Describe the rationale for the review in the context of existing knowledge. | p2 |
| Objectives | 4 | Provide an explicit statement of the objective(s) or question(s) the review addresses. | p2 |
| **METHODS** |  |  |  |
| Eligibility criteria | 5 | Specify the inclusion and exclusion criteria for the review and how studies were grouped for the syntheses. | p3 |
| Information sources | 6 | Specify all databases, registers, websites, organisations, reference lists and other sources searched or consulted to identify studies. Specify the date when each source was last searched or consulted. | P4 |
| Search strategy | 7 | Present the full search strategies for all databases, registers and websites, including any filters and limits used. | Protocol |
| Selection process | 8 | Specify the methods used to decide whether a study met the inclusion criteria of the review, including how many reviewers screened each record and each report retrieved, whether they worked independently, and if applicable, details of automation tools used in the process. | p5 |
| Data collection process | 9 | Specify the methods used to collect data from reports, including how many reviewers collected data from each report, whether they worked independently, any processes for obtaining or confirming data from study investigators, and if applicable, details of automation tools used in the process. | p5 |
| Data items | 10a | List and define all outcomes for which data were sought. Specify whether all results that were compatible with each outcome domain in each study were sought (e.g. for all measures, time points, analyses), and if not, the methods used to decide which results to collect. | p5 |
|  | 10b | List and define all other variables for which data were sought (e.g. participant and intervention characteristics, funding sources). Describe any assumptions made about any missing or unclear information. | N/A |
| Study risk of bias assessment | 11 | Specify the methods used to assess risk of bias in the included studies, including details of the tool(s) used, how many reviewers assessed each study and whether they worked independently, and if applicable, details of automation tools used in the process. | p7 |
| Effect measures | 12 | Specify for each outcome the effect measure(s) (e.g. risk ratio, mean difference) used in the synthesis or presentation of results. | N/A |
| Synthesis methods | 13a | Describe the processes used to decide which studies were eligible for each synthesis (e.g. tabulating the study intervention characteristics and comparing against the planned groups for each synthesis (item #5)). | p5 |
|  | 13b | Describe any methods required to prepare the data for presentation or synthesis, such as handling of missing summary statistics, or data conversions. | Table 1. p4 |
|  | 13c | Describe any methods used to tabulate or visually display results of individual studies and syntheses. | N/A |
|  | 13d | Describe any methods used to synthesize results and provide a rationale for the choice(s). If meta-analysis was performed, describe the model(s), method(s) to identify the presence and extent of statistical heterogeneity, and software package(s) used. | p8 |
|  | 13e | Describe any methods used to explore possible causes of heterogeneity among study results (e.g. subgroup analysis, meta-regression). | N/A |
|  | 13f | Describe any sensitivity analyses conducted to assess robustness of the synthesized results. | N/A |
| Reporting bias assessment | 14 | Describe any methods used to assess risk of bias due to missing results in a synthesis (arising from reporting biases). | N/A |
| Certainty assessment | 15 | Describe any methods used to assess certainty (or confidence) in the body of evidence for an outcome. | N/A |
| **RESULTS** |  |  |  |
| Study selection | 16a | Describe the results of the search and selection process, from the number of records identified in the search to the number of studies included in the review, ideally using a flow diagram. | Figure 1. p7 |
|  | 16b | Cite studies that might appear to meet the inclusion criteria, but which were excluded, and explain why they were excluded. | Figure 1. p7 |
| Study characteristics | 17 | Cite each included study and present its characteristics. | Table 3. p13 |
| Risk of bias in studies | 18 | Present assessments of risk of bias for each included study. | Table 3. p13 |
| Results of individual studies | 19 | For all outcomes, present, for each study: (a) summary statistics for each group (where appropriate) and (b) an effect estimate and its precision (e.g. confidence/credible interval), ideally using structured tables or plots. | Table 3. p13 |
| Results of syntheses | 20a | For each synthesis, briefly summarise the characteristics and risk of bias among contributing studies. | p9-12 & p19-21 |
|  | 20b | Present results of all statistical syntheses conducted. If meta-analysis was done, present for each the summary estimate and its precision (e.g. confidence/credible interval) and measures of statistical heterogeneity. If comparing groups, describe the direction of the effect. | N/A |
|  | 20c | Present results of all investigations of possible causes of heterogeneity among study results. | N/A |
|  | 20d | Present results of all sensitivity analyses conducted to assess the robustness of the synthesized results. | N/A |
| Reporting biases | 21 | Present assessments of risk of bias due to missing results (arising from reporting biases) for each synthesis assessed. | N/A |
| Certainty of evidence | 22 | Present assessments of certainty (or confidence) in the body of evidence for each outcome assessed. | N/A |
| **DISCUSSION** |  |  |  |
| Discussion | 23a | Provide a general interpretation of the results in the context of other evidence. | p20 |
|  | 23b | Discuss any limitations of the evidence included in the review. | p24 |
|  | 23c | Discuss any limitations of the review processes used. | p24 |
|  | 23d | Discuss implications of the results for practice, policy, and future research. | p25 |
| **OTHER INFORMATION** |  |  |  |
| Registration and protocol | 24a | Provide registration information for the review, including register name and registration number, or state that the review was not registered. | p5 |
|  | 24b | Indicate where the review protocol can be accessed, or state that a protocol was not prepared. | p5 |
|  | 24c | Describe and explain any amendments to information provided at registration or in the protocol. | N/A |
| Support | 25 | Describe sources of financial or non-financial support for the review, and the role of the funders or sponsors in the review. | p26 |
| Competing interests | 26 | Declare any competing interests of review authors. | p26 |
| Availability of data, code and other materials | 27 | Report which of the following are publicly available and where they can be found: template data collection forms; data extracted from included studies; data used for all analyses; analytic code; any other materials used in the review. | N/A |

